# Patterns of resistance to antibiotics of *Enterococcus faecalis* in clinical isolates of diabetic foot and presence of culture resistant to linezolid

**DOI:** 10.1101/2020.11.09.20228478

**Authors:** Jorge Angel Almeida Villegas, Luis Enrique García Fernández, Iris Mellolzy Estrada Carrillo, Harold Mondragon Reyes, Mariana Aguilar Sánchez, Silvia Patricia Peña

## Abstract

**Introduction:** High levels of hyperglycemia lead to diabetes, the same levels that if not controlled increase the risk of diabetic neuropathy, which over time translates into loss of sensation and lesions that progress and lead to diabetic foot may occur, microorganisms Mainly Gram positive cocci and Gram negative bacilli, as well as yeasts are those that are mainly found in this metabolic and infectious pathology.

**Methods:** Diabetic foot wounds were studied in patients aged 45-54 years, with a mean age of 50 years, who had different periods of time with a hyperglycemia problem and different complications associated with diabetic foot, and were not considered as exclusion criteria. With a total of 41 cultures, 25 belong to male patients and 16 to female patients. All of these in the Toluca Valley, Mexico. The primary swab was reseeded in chromogenic agar, BHI, Salt and mannitol, calf blood, EMB and MacConkey, after 6 hours after taking the sample. In addition to microbial reseeding, a Gram stain was performed on each of the samples. The Petri dishes were placed in an incubation oven for 18 hours at 35 ° C with ± 2 ° C. Bacterial identification was performed on automated equipment from Beckman Coulter, as well as antibiotic sensitivity tests.

**Results:** 35 positive cultures were obtained, of which they had a single microbial agent and some cases with two agents, which could be bacteria-bacteria or bacteria-yeast. With 14 positive strains for *Enterococcus faecalis*, with 100% sensitivity for cell wall inhibitors, and high resistance to tetracycline with 85.71% and 92.86% resistance to the macrolide erythromycin. In addition, there is a strain that was resistant to linezolid, and variable resistance patterns in fluoroquinolones and other antibiotics.

## Introduction

Diabetes is a disease in which there is a disorder in the metabolism caused by hyperglycemia due to a deficiency or insufficiency of insulin. Diabetes mellitus in Mexico is one of the main chronic and degenerative diseases that, if not properly treated, lead to serious systemic complications such as kidney damage or vascular insufficiencies. These complications generate greater pressure between the muscle and the nerves that connect with the peripheral nervous system with somatic action and, together with high glucose levels, lead the body to diabetic peripheral neuropathy, which is why sensitivity to pain is reduced.^3-6^ Together with venous insufficiency they create macro and micro vasculopathies causing ulcers in the foot and adding that the body is immunocompromised due to the imbalance in homeostasis they facilitate that foot wounds are perfect for microbial development, being opportunistic microorganisms the cause of diseases infectious diseases in the diabetic foot, which in the worst case can lead to amputation of the limb or an infectious systemic disease. ^2,4,9^ Among the most frequent microorganisms in this type of wound are the enterobacteria with E.coli, Citrobacter and Klebsiella or ESKAPE group, in the same way it is very common to find the genus Staphy lococcus, due to immunosuppression, the bacteria of the skin microbiota also cause opportunistic infections, poor hygiene causes the Enterococcus genus to be isolated a lot, and one of the bacteria that is also isolated due to being in contact with moisture is the genus Pseudomonas, although yeasts of the genus Candida are also more frequently isolated.^9,10^

One of the complications of diabetic foot infections is that most of these bacteria have created a multi-resistance to several antibiotics, in addition to the fact that the patient does not always take the treatment properly, which ends up causing an amputation in 70% of patients. emergency cases in hospitals and of that 70%, approximately 20% end up in sepsis. Therefore, a good microbiological diagnosis is very important in addition to the study of the sensitivity towards first, second and third and fourth generation antibiotics to avoid said bacterial resistance which is a problem in developing countries such as Mexico due to treating these infections with treatments. 4th generation empirics. Many specialists and doctors believe in the fallacy that 1st and 2nd generation antibiotics no longer have an effect on the main pathogenic bacteria, it is known that nosocomial bacteria have created a multi-resistance, but in patients with diabetic foot infection, many of these bacteria Opportunists and pathogens have not yet created this multi-resistance, so they can still be treated with penicillin or Trimethoprim sulfamethoxazole as the first option in treatment. At an epidemiological level, health specialists must also know which is the main isolated microorganism to be able to search for new alternatives to the prevention of an infection in a diabetic foot, of course, preventing diabetes is better than preventing its complications, but once the disease is diagnosed. We can control diabetes and prevent opportunistic infections, many of these opportunistic bacteria are due to poor hygiene on the part of the patient, so it can be prevented by modifying or adding specifications in personal hygiene care by medical indications or specialists of health.^1,7^

## Methods

Diabetic foot wounds were studied in patients aged 45-54 years, with a mean age of 50 years, who had different periods of time with a hyperglycemia problem and different complications associated with diabetic foot, and were not considered as exclusion criteria. With a total of 41 cultures, 25 belong to male patients and 16 to female patients. All of these in the Toluca Valley, Mexico

To collect the microbiology sample, a sterile swab and Copan® transport medium were used, the swab was taken from the area with the greatest suppuration in the diabetic foot lesion.

The primary swab was reseeded in chromogenic agar, BHI, Salt and mannitol, calf blood, EMB and MacConkey, after 6 hours after taking the sample. In addition to microbial reseeding, a Gram stain was performed on each of the samples.

The Petri dishes were placed in an incubation oven for 18 hours at 35 ° C with ± 2 ° C. Subsequently, some patterns were identified with the used media, the chromogenic agar colonies were re-suspended in medium to dilute for automated equipment, with a standard McFarland scale. The growth in calf blood agar culture medium was used to count CFU’s.

Bacterial identification was performed on automated equipment from Beckman Coulter, as well as antibiotic sensitivity tests.

## Results and discussion

Of the 41 cultures, 35 were positive (85.36%) and 6 negative for microbial development (14.64%). 5 cultures (14.28%) of the positive cultures presented poly-microbial infection, with at least two bacteria or the presence of bacteria and yeast.

14 *Enterococcus faecalis* strains were isolated from the 41 cultures, which no clinical sample showed the presence of bacterial or yeast co-infection. Obtaining the resistance patterns shown in table number 1.

**Table 1.** Antibiotic resistance patterns, where the letter S = sensitive, R = resistant and I intermediate sensitivity.

|  |  |  |  |  |  |  |  |  |  |  |  |  |  |  |  |
| --- | --- | --- | --- | --- | --- | --- | --- | --- | --- | --- | --- | --- | --- | --- | --- |
| AMPICILLIN | S | S | S | S | S | S | S | S | S | S | S | S | S | S | 14 S |
| CIPROFLOXACIN | S | R | R | R | S | S | S | R | R | R | S | R | R | R | 9R/ 5S |
| DAPTOMYCIN | S | S | S | S | S | S | S | S | S | S | S | S | S | S | 14 S |
| ERYTHROMYCIN | R | R | R | R | S | R | R | R | R | R | R | R | R | R | 13R/ 1S |
| LEVOFLOXACIN | S | R | R | R | S | S | S | R | R | R | S | R | R | R | 9R/ 5S |
| LINEZOLID | S | S | S | R | S | S | S | S | S | S | S | S | S | S | 13S/ 1R |
| PENICILLIN | S | S | S | S | S | S | S | S | S | S | S | S | S | S | 14 S |
| RIFAMPICIN | S | R | S | R | R | S | R | R | R | S | R | R | S | I | 5S, 1I, 8R |
| TETRACYCLINE | R | R | R | R | S | R | R | R | R | S | R | R | R | R | 2S/ 12 R |
| VANCOMYCIN | S | S | S | S | S | S | S | S | S | S | S | S | S | S | 14 S |

Of which the 14 strains show 100% sensitivity for the antibiotics ampicillin, penicillin, vancomycin and daptomycin respectively. This indicates that all the antibiotics that interfere with the synthesis of the bacterial cell wall such as the beta-lactams ampicillin and penicillin, and other synthesis inhibitors such as vancomycin and daptomycin are still effective. In the case of the fluoroquinolones ciprofloxacin and levofloxacin, 9 strains are resistant to both antibiotics respectively (64.28%) and 5 are sensitive to the same antibiotics (35.72%), for rifampicin it was found that 8 strains (57.14%) are resistant, 1 (7.14%) presents intermediate sensitivity and 5 show sensitivity (35.72%), the antibiotic tetracycline was obtained 12 strains with resistance (85.71%) and only 2 are sensitive strains (14.29%), the antibiotic to which the greatest resistance was had is to Erythromycin macrolide with 13 resistant strains (92.86%) and only 1 sensitive strain (7.14%). However, an important finding is the presence of a strain that showed resistance to linezolid, which despite being 7.14%, the presence of resistance to this antibiotic suggests a problem, which can lead to multiple resistance. Graph 1 shows resistance patterns for some classes of antibiotics, highlighting erythromycin and tetracycline as the drugs with the highest resistance by Enterococcus faecalis.

**Graph 1.**
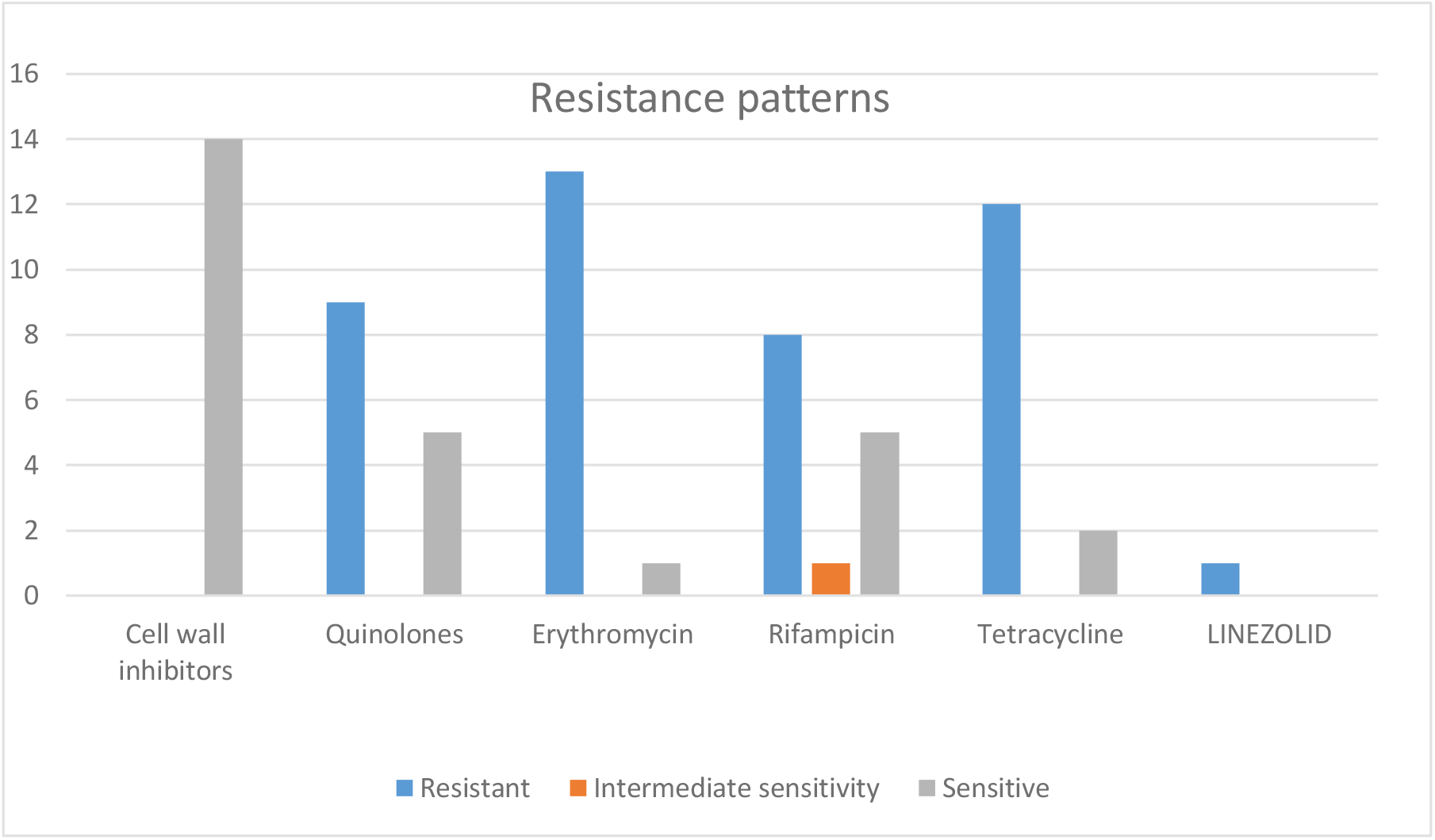
Patterns of resistance to some classifications of antibiotics in *Enterococcus faecalis*.

## Conclusion

In the present research work, the isolation and identification of antibacterial resistance patterns by the bacterium *Enterococcus faecalis* was carried out, where it is shown that the antibiotics tetracillin with 85.71% and the macrolide erythromycin 92.86% are those with the highest resistance due to part of the isolated strains, while the inhibitors of cell wall formation penicillin and ampicillin, beta-lactam antibiotics, as well as daptomycin and the glucopeptide vancomycin, show a sensitivity of 100%, which suggests that the enteroccous faecalis strains in the Toluca can still be treated with these drugs that present the first line of choice. One more fact that we highlight is that although it is true that the bacteria are common in diabetic foot infections, the number of isolates in this study is high with 34.14% of the total isolated bacteria. Suggesting the infection appeared in these patients due to poor hygiene. Something that is alarming in the study is the presence of a strain resistant to linezolid, which is a drug used to treat infections with Gram-positive bacteria when vancomycin does not represent a solution.

## Data Availability

All readers can make use of the information included in the work

## Code of ethics

The ethics committee of the Microtec laboratory approved the implementation of the following project, the same research work that does not violate or violate the security and identity of individuals since samples that are considered routine were collected.

## Financing

The following research work does not receive funding for its preparation

## Gratefulness

To all the people who provided a sample to carry out the project, as well as to all the institutions that allowed obtaining the taking and carrying out of the experiments. The following work is dedicated especially to Victoria, daughter of the main autor

